# Antenatal Screening for Sexually Transmitted Infections to Reduce Preterm Birth or Low Birthweight (Philani Ndiphile Study): A Randomized Three-Group Trial

**DOI:** 10.64898/2026.04.15.26350805

**Authors:** Andrew Medina-Marino, Chibuzor M Babalola, Mandisa M Mdingi, Melissa L Wilson, Freedom Mukomana, Christina A Muzny, Christopher M Taylor, Ranjana MS Gigi, Hyunsul Jung, Nicola Low, Remco PH Peters, Jeffrey D Klausner

**Author notes:** co-Lead Authors (Andrew Medina-Marino, Chibuzor M Babalola).

## Abstract

**Background:** *Chlamydia trachomatis*, *Neisseria gonorrhoeae*, and *Trichomonas vaginalis* are curable sexually transmitted infections (STIs) associated with adverse birth outcomes. Most infections are asymptomatic. Whether antenatal STI screening improves birth outcomes remains uncertain.

**Methods:** In a randomized three-group trial in South Africa, pregnant women aged 18 years or older were assigned before 27 weeks’ gestation to: (1) screening and treatment for *Chlamydia trachomatis, Neisseria gonorrhoeae*, and *Trichomonas vaginalis* at enrollment, with tests-of-cure (One-Time Screening); (2) screening and treatment at enrollment, repeated at 30 to 34 weeks (Two-Time Screening); or (3) Standard-of-Care (Syndromic management). The primary outcome was a composite of preterm birth (<37 weeks’ gestation) or low birthweight (<2500 g), analyzed in the modified intention-to-treat population of participants with live births. Components of the composite outcome were evaluated individually as the main secondary outcomes. The study was registered with ClinicalTrials.gov, NCT04446611.

**Findings:** Of 2247 enrolled participants, 1910 had live births. The composite outcome occurred in 22·9% of the One-Time Screening group (risk ratio [RR] 0·99; 95% confidence interval [CI] 0·81–1·21), 20·6% of the Two-Time Screening group (RR 0·89; 95% CI 0·72–1·09), compared with 23·2% of the Standard-of-Care group. Preterm birth occurred in 18·9% of the One-Time Screening group (RR 1·00; 95% CI 0·80–1·26), 14·5% of the Two-Time Screening group (RR 0·77; 95% CI 0·60–0·99), and 18·8% of the Standard-of-Care group. Low birthweight occurred in 14·1% of the One-Time Screening group (RR 1·10; 95% CI 0·83–1·46), 12·9% of the Two-Time Screening group (RR 1·01; 95% CI 0·76–1·35), and 12·8% of the Standard-of-Care group.

**Interpretation:** Neither screening strategy for *Chlamydia trachomatis*, *Neisseria gonorrhoeae*, and *Trichomonas vaginalis* reduced the primary composite outcome of preterm birth or low birthweight, or low birthweight alone. The Two-Time antenatal STI screening strategy, however, reduced preterm birth by 23%.

## INTRODUCTION

Curable sexually transmitted infections (STIs)—including *Chlamydia trachomatis*, *Neisseria gonorrhoeae*, and *Trichomonas vaginalis*—are associated with adverse birth outcomes, including preterm birth and low birthweight.^1–3^ The burden of STIs is highest in low- and middle-income countries, with their combined prevalence exceeding 30% among pregnant women in sub-Saharan Africa.^4,5^ The World Health Organization (WHO) recommends syndromic management of symptomatic women during antenatal care in resource-limited settings, leaving asymptomatic infections untreated.^6,7^

Point-of-care molecular diagnostics have made screening and treatment increasingly feasible.^8^ However, evidence of its effect on pregnancy outcomes remains limited, with no definitive randomized controlled trial evidence from sub-Saharan Africa to inform regional policy.^9^ In response, the WHO and global health commissions have prioritized evaluation of antenatal STI screening and treatment, issuing a conditional recommendation in 2025 to screen and treat where resources permit.^10,11^ Existing interventional studies have yielded mixed results. An early trial of presumptive antenatal antibiotic treatment Uganda reported reductions in adverse birth outcomes, whereas trials in Malawi (presumptive azithromycin) and the United States (*C. trachomatis* treatment) found no overall benefit.^12–14^

To date, only one large, randomized trial has directly evaluated antenatal STI screening and treatment: the WANTAIM cluster-randomized trial in Papua New Guinea. ^15^ WANTAIM assessed three-time antenatal screening in a low-HIV, malaria-endemic setting between 2017 and 2022 and did not demonstrate an overall reduction in preterm birth or low birthweight compared with syndromic management, although benefit was observed among women infected with *N. gonorrhoeae* at enrollment.^15^ Post-hoc analyses from a smaller non-randomized study in Botswana (MADUO study) hypothesized potential benefit of antenatal STI screening and treatment to reduce preterm or low birthweight among nulliparous women.^16^ Collectively, these findings highlight that evidence for antenatal STI screening remains heterogeneous and potentially context dependent, and that definitive evidence on antenatal screening strategies is still lacking.

Therefore, we undertook the Philani Ndiphile trial (“Philani,” meaning “healthy mother, healthy baby” in isiXhosa) in South Africa, a high STI and HIV prevalence setting, to evaluate the effectiveness of two different antenatal STI screening strategies on adverse birth outcomes.

## METHODS

### Study Design and Participants

Philani was a three-group, individually randomized, open-label clinical trial conducted at four government health clinics in Buffalo City Metropolitan District, Eastern Cape, South Africa.^17^ The trial was registered at ClinicalTrials.gov (NCT04446611) and is reported in accordance with the CONSORT 2025 Statement.^18^ Eligible participants were pregnant women aged ≥18 years, presenting for their first antenatal visit and intending to remain through delivery; excluded if they were unable to or declined consent.

Enrollment occurred from March 29, 2021, through May 31, 2024, with follow-up for pregnancy outcome collection completed through February 2025. In November 2021, the gestational age eligibility threshold was expanded from <20 to <27 weeks, after 328 participants (14·6%) had been enrolled, to mitigate COVID-19 delays and align with similar studies.^15,16^ Participants were randomly assigned (1:1:1) to: (1) One-Time Screening — screening and treatment for *C. trachomatis, N. gonorrhoeae, and T. vaginalis* at first visit and, if positive, a test-of-cure; (2) Two-Time Screening, — screening and treatment at first visit, and a routine third-trimester visit between 30 and 34 weeks’ (no tests of cure); or (3) Standard-of-Care — syndromic management in women presenting with vaginal discharge syndrome^19^

### Trial Oversight

The study protocol was approved by the University of Cape Town Faculty of Health Sciences Human Research Ethics Committee (reference 676/2019) and the Eastern Cape Provincial Research Committee, with reliance agreements from collaborating institutions. All participants provided written informed consent. An independent Data and Safety monitoring board (DSMB) reviewed study conduct every 6 months. The final analysis was performed on a locked database, and the DSMB was unblinded to the results till completion of the primary statistical analysis.

### Randomization

Participants were randomly assigned in blocks of 15, stratified by clinic, to one of three groups using a computer-generated allocation sequence by a data management specialist not directly involved in study conduct. Clinic teams were unaware of block size to minimize potential recruitment bias. Outcome assessors and data analysts were masked to group assignment.

### Procedures

#### Baseline

At enrollment (first antenatal visit), participants were informed of their group assignment after providing consent. All received routine antenatal care in accordance with national guidelines,^19^ including HIV and syphilis testing. A transabdominal ultrasound was performed to confirm gestational age and pregnancy viability, factoring a composite of transabdominal biometric measurements—including biparietal diameter, head circumference, abdominal circumference, and femur length to ensure dating accuracy. Demographic and clinical data were collected by trained study nurses. Clinician-collected vaginal swabs were obtained during routine antenatal clinical examinations.

Participants in the One-Time and Two-Time Screening Groups were tested at enrollment for *C. trachomatis, N. gonorrhoeae, and T. vaginalis* using the GeneXpert® CT/NG and TV assays, Cepheid, USA at point-of-care. Women with positive results received treatment at the same visit or were actively recalled within one week of results. Treatment included azithromycin (1 g orally) for *C. trachomatis*, ceftriaxone (500 mg intramuscularly) for *N. gonorrhoeae*, and metronidazole (400 mg orally twice daily for 7 days) for *T. vaginalis*. ^20^ Single-dose medications and the first dose of metronidazole were directly observed.

Participants in the Standard-of-Care Group did not receive real time antenatal STI testing; vaginal swabs were collected and stored for post-trial testing. Women presenting with vaginal discharge syndrome—abnormal vaginal discharge with or without other urogenital symptoms—received syndromic management (presumptive single dose azithromycin, ceftriaxone, and metronidazole (2 g)), consistent with national guidelines.^19^

Symptomatic participants in both screening groups who tested negative for all three STIs received 2 g metronidazole only (i.e., as presumptive bacterial vaginosis treatment), according to current practice at study sites. All treated for an STI—whether through aetiologic screening or syndromic management—received a notification slip to encourage their partner(s) to seek treatment (standard practice).

#### Antenatal follow-up

In the One-Time Screening Group, participants who tested positive at enrollment were scheduled for a test-of-cure 3 to 5 weeks later and re-treated if still positive. Test-of-cure was targeted to the index infection treated; incident *C. trachomatis* or *N. gonorrhoeae* infections detected with the combined CT/NG assay were also treated. In the Two-Time Screening Group, screening and treatment were repeated at a third trimester antenatal visit between 30- and 34-weeks’ gestation (no tests-of-cure).

#### Postnatal follow-up

A postnatal visit was scheduled within two weeks of delivery to retrieve birth outcomes. Additionally, all participants, including those in the Standard-of-Care Group, underwent testing at the postnatal visit for *C. trachomatis, N. gonorrhoeae, and T. vaginalis* and treatment if positive. No STI testing was conducted beyond this 2-week window. Birth outcomes were ascertained from hand-held maternal discharge summaries during an in-person 2-week postnatal visit or, for participants captured later, at the routine 6-week infant immunization visit. Participant-held records were supplemented with facility data. Adverse Events were defined per protocol consistent with University of Cape Town Human Research Ethics Committee Standard Operating Procedures and reviewed routinely by the DSMB. Specifically, serious adverse events encompassed perinatal events resulting in maternal or newborn death, life-threatening illness, hospitalization, disability, or congenital abnormality. Study data were captured electronically into a password-protected, Health Insurance Portability and Accountability Act, and Protection of Personal Information Act-compliant Research Electronic Data Capture database.^21,22^

### Study Outcomes

The primary outcome was a composite of preterm birth (<37 weeks’ gestation) or low birthweight (<2500 g), estimated based on the ultrasound-dating at enrollment. Measurement of infant weight at birth is a standard clinical procedure in South African public-sector obstetric care, using calibrated facility scales.. The components of the composite—preterm birth and low birthweight—were evaluated individually as the main secondary outcomes for this manuscript. A prespecified subgroup analysis among participants with laboratory-confirmed STIs at baseline was also conducted, given its direct relevance to the intervention and for comparability with other trials. The STI positivity at each study visit (antenatal and postnatal) is presented descriptively to provide context on infection patterns over the pregnancy course. Other planned secondary and exploratory outcomes included severe subtypes of preterm birth and low birthweight (moderate, very, or extremely preterm), pregnancy loss, and hypertensive disorders. These will be presented in subsequent study reports.

### Statistical Analyses

We hypothesized that either of the one time or two-time antenatal screening and treatment strategies for *C. trachomatis, N. gonorrhoeae, or T. vaginalis* — would reduce preterm birth or low birthweight.

Assuming 23% baseline incidence and a 25% relative reduction, an effective sample of 2079 women with live births (693 per group) would provide 80% power at α = 0·05. Calculations were performed using PASS 2023 (NCSS, LLC, Kaysville, UT, USA). Enrollment concluded in May 2024 at 2,247 participants due to funding constraints. Prospective ascertainment of pregnancy outcomes was completed in February 2025, available for 2,119 participants (94% of enrolled).

Main outcome analyses were conducted in the modified intention-to-treat population, defined as randomized participants with a documented live birth and available outcome data, with the participant as the unit of analysis, regardless of attendance at follow-up STI testing visits (test-of-cure or third trimester screening). In thirty-five participants with multiple gestations, the outcome was defined at the maternal pregnancy level if either infant was preterm or low birthweight or both. Risks were calculated as the proportion of participants within each randomized group who experienced the outcome. Risk ratios and 95% confidence intervals were estimated using binomial regression (with a log link), comparing each screening group to Standard-of-Care. Continuous data are presented as medians with interquartile ranges, and categorical variables as frequencies with percentages. Statistical analyses were conducted by two separate analysts not directly involved in the study, using Stata (version 17; StataCorp, College Station, TX, USA) and R (version 4.4.1; R Foundation for Statistical Computing, Vienna, Austria). Complete-case data were used; missing values were not imputed given the low proportion of missingness. As prespecified in the statistical analysis plan, if predictors with strong prognostic relevance to the primary outcome appeared unbalanced between groups at baseline, they would be considered for inclusion in the primary regression models..

## RESULTS

Of 2940 women assessed for eligibility, 2247 were enrolled and randomly assigned to the One-Time Screening (n=754, 33·6%), the Two-Time Screening (n=738, 32·8%), or Standard-of-Care (n=755, 33·6%) (**Figure 1**). Pregnancy outcome data were retrieved for 2119 participants (94·3%) before study closure. Among those, 1910 had live births and comprised the modified intention-to-treat population: 625 in the One-Time Screening Group, 642 in the Two-Time Screening Group, and 643 in the Standard-of-Care Group.

**Figure 1.**
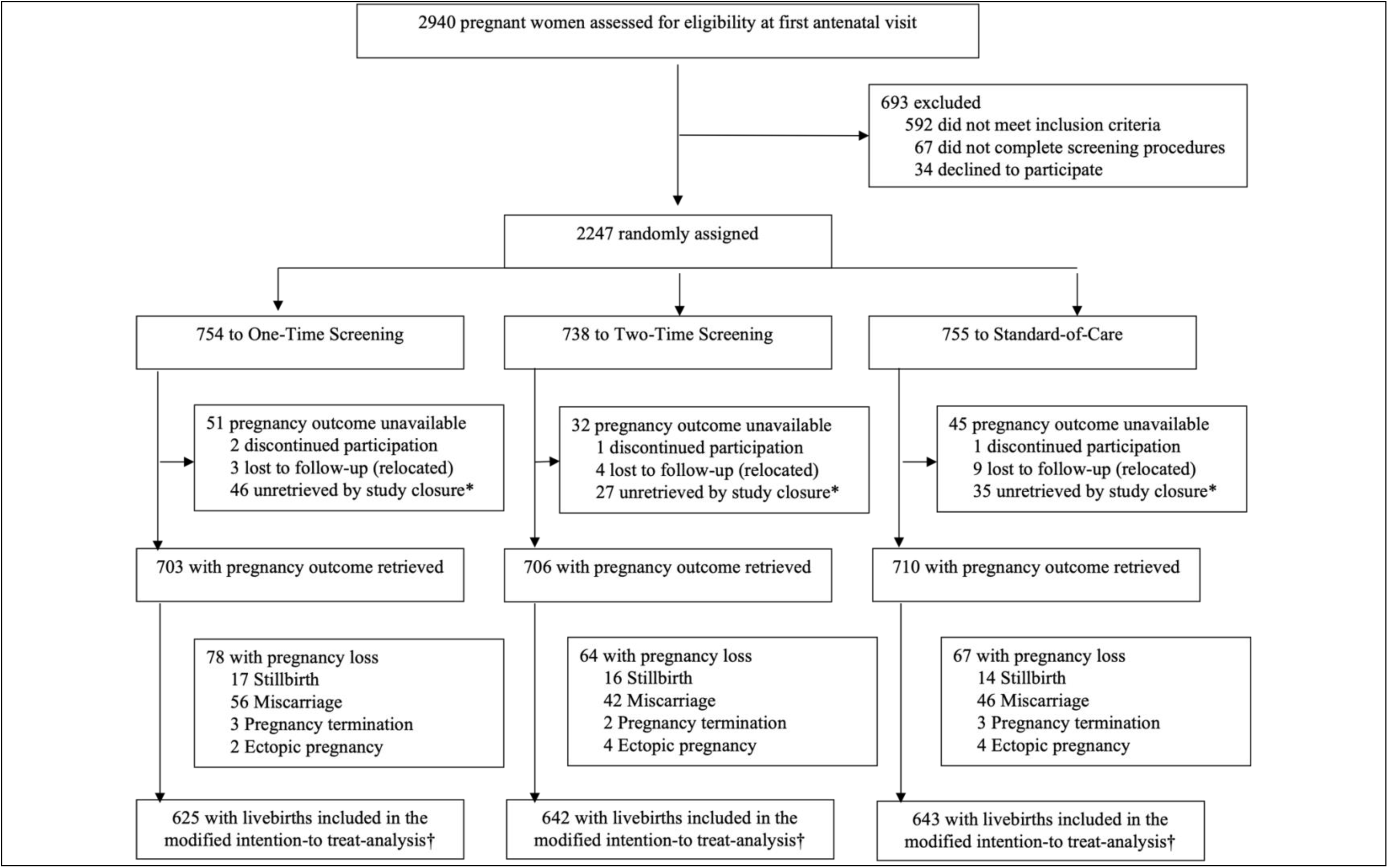
Trial Profile. *Dropouts from participants whose outcomes could not be ascertained before funding period elapsed in March 2025 †1910 total participants with a live-birth delivery, including 35 with twin gestations, analyzed for main endpoints (preterm or low birthweight)

### Baseline Characteristics

Across all study groups, the median gestational age was 13 weeks (IQR 8–18) at the first antenatal visit and 30 weeks (IQR 30–32) at the routine third-trimester visit (**Table 1**) where only the Two-Time Screening group received repeat STI screening and treatment. Among participants treated for STI in the One-Time Screening group, the median gestational age at the test-of-cure visit was 16 weeks (IQR 11–23).

**Table 1.** Baseline Characteristics of Enrolled Participants.

| **Table 1. Baseline Characteristics of Enrolled Participants** |  |  |  |  |
| --- | --- | --- | --- | --- |
|  | Total Enrolled | One-Time Screening | Two-Time Screening | Standard of Care |
|  | N = 2247 | N= 754 | N=738 | N=755 |
| Antenatal Enrollment and Follow-up Timing (in weeks) * |  |  |  |  |
| Gestational age at the first antenatal visit |  |  |  |  |
| Median (IQR) | 13 (8–18) | 13 (8–18) | 14 (9 –18) | 13 (8 –18) |
| Gestational age at the test-of-cure visit |  |  |  |  |
| Median (IQR) | — | 16 (11–23) | Not applicable | Not applicable |
| Gestational age at the third trimester visit |  |  |  |  |
| Median (IQR) | 30 (30–32) | 30 (30–32) | 30 (30–32) | 30 (30–32) |
| Socio-demographics |  |  |  |  |
| Age, years |  |  |  |  |
| Median (IQR) | 28 (24–33) | 28 (24–33) | 28 (23–33) | 28 (24–33) |
| Highest level of education |  |  |  |  |
| High school or lower | 1956 (87·1%) | 654 (86·9%) | 651 (88·2%) | 651 (86·3%) |
| Tertiary Education | 289 (12·9%) | 99 (13·1%) | 87 (11·8%) | 103 (13·7%) |
| Missing | 2 | 1 | 0 | 1 |
| Personal monthly income† |  |  |  |  |
| No personal income | 1289 (57·4%) | 421 (55·8%) | 437 (59·2%) | 431 (57·1%) |
| ≤10,000 ZAR | 907 (40·3%) | 315 (41·8%) | 288 (39·0%) | 304 (40·2%) |
| >10,000 ZAR | 51 (2·3%) | 18 (2·4%) | 13 (1·8%) | 20 (2·6%) |
| Employment status |  |  |  |  |
| Not employed | 1281 (57·0%) | 417 (55·3%) | 436 (59·1%) | 428 (56·7%) |
| Employed, including self-employment | 966 (43·0%) | 337 (44·7%) | 302 (40·9%) | 327 (43·3%) |
| Sexual and Socio-behavioral History |  |  |  |  |
| Relationship with current sex partner |  |  |  |  |
| No current partner | 64 (2·8%) | 18 (2·4%) | 23 (3·1%) | 23 (3·0%) |
| Married or steady relationship | 2092 (93·1%) | 710 (94·2%) | 676 (91·6%) | 706 (93·5%) |
| In casual relationship(s) | 91 (4.1%) | 26 (3.4%) | 39 (5.3%) | 26 (3.4%) |
| Partner may have other sex partners |  |  |  |  |
| Participant does not know | 344 (16.1%) | 121 (16.8%) | 96 (13.8%) | 127 (17.7%) |
| No | 1177 (55.2%) | 415 (57.7%) | 384 (55.2%) | 378 (52.8%) |
| Yes | 610 (28.6%) | 183 (25.5%) | 216 (31.0%) | 211 (29.5%) |
| Missing | 116 | 35 | 42 | 39 |
| Alcohol consumption in this pregnancy |  |  |  |  |
| No | 1614 (71.8%) | 558 (74.0%) | 509 (69.0%) | 547 (72.5%) |
| Yes | 633 (28.2%) | 196 (26.0%) | 229 (31.0%) | 208 (27.5%) |
| Smokes cigarettes† |  |  |  |  |
| No | 1538 (91.2%) | 519 (91.9%) | 492 (89.0%) | 527 (92.8%) |
| Yes | 148 (8.8%) | 46 (8.1%) | 61 (11.0%) | 41 (7.2%) |
| Missing | 561 | 189 | 185 | 187 |
| Obstetric History§ |  |  |  |  |
| Nulliparous |  |  |  |  |
| No | 1442 (64.2%) | 482 (63.9%) | 489 (66.3%) | 471 (62.4%) |
| Yes, first time pregnant | 673 (30.0%) | 225 (29.8%) | 211 (28.6%) | 237 (31.4%) |
| Yes, previous pregnancy ended in a loss | 132 (5.9%) | 47 (6.2%) | 38 (5.1%) | 47 (6.2%) |
| Previous Pregnancy Loss§ | (n = 1574) | (n = 529) | (n = 527) | (n = 518) |
| No | 1071 (69.9%) | 350 (68.0%) | 367 (71.5%) | 354 (70.1%) |
| Yes | 462 (30.1%) | 165 (32.0%) | 146 (28.5%) | 151 (29.9%) |
| Missing | 41 | 14 | 14 | 13 |
| Previous Preterm Birth§ | (n = 1401) | (n = 468) | (n = 475) | (n = 458) |
| No | 1212 (87.2%) | 403 (87.2%) | 426 (90.1%) | 383 (84.2%) |
| Yes | 178 (12.8%) | 59 (12.8%) | 47 (9.9%) | 72 (15.9%) |
| Missing | 11 | 6 | 2 | 3 |
| Sexually Transmitted Infection (STI) or Reproductive Tract Infection at Enrollment |  |  |  |  |
| *C. trachomatis*, *N. gonorrhoeae*, or *T. vaginalis* infection Status ¶ |  |  |  |  |
| Uninfected | 1642 (73.3%) | 561 (74.4%) | 552 (74.8%) | 529 (70.8%) |
| Tested positive for any of the three STIs | 597 (26.7%) | 193 (25.6%) | 186(25.2%) | 218 (29.2%) |
| *C. trachomatis* | 353 (15.8%) | 112 (14.9%) | 107 (14.5%) | 134 (17.9%) |
| *N. gonorrhoeae* | 123 (5.5%) | 37 (4.9%) | 36 (4.9%) | 50 (6.7%) |
| *T. vaginalis* | 235 (10.5%) | 81 (10.7%) | 71 (9.6%) | 83 (11.1%) |
| Missing | 8 | 0 | 0 | 8 |
| Presented with Vaginal Discharge Syndrome |  |  |  |  |
| No | 1929 (85.8%) | 647 (85.8%) | 640 (86.7%) | 642 (85.0%) |
| Yes | 318 (14.2%) | 107 (14.2%) | 98 (13.3%) | 113 (15.0%) |
| Syphilis Rapid Diagnostic Test Results |  |  |  |  |
| Negative | 2160 (96.3%) | 728 (96.7%) | 709 (96.2%) | 723 (95.9%) |
| Positive | 84 (3.7%) | 25 (3.3%) | 28 (3.8%) | 31 (4.1%) |
| Missing | 3 | 1 | 1 | 1 |
| Bacterial vaginosis (Nugent ≥ 7) | | | | |
| No | 988 (48.3%) | 335 (49.2%) | 315 (46.7%) | 338 (49.1%) |
| Yes | 1056 (51.7%) | 346 (50.8%) | 359 (53.3%) | 351 (50.9%) |
| Missing | 203 | 73 | 64 | 66 |
| HIV infection status |  |  |  |  |
| Uninfected | 1592 (70.9%) | 529 (70.2%) | 515 (69.8%) | 548 (72.6%) |
| Living with HIV | 655 (29.1%) | 225 (29.8%) | 223 (30.2%) | 207 (27.4%) |
| Already on antiretroviral therapy | 535/655 (81.7%) | 193/225 (85.7%) | 179/223 (80.3%) | 163/207 (78.7%) |
| HIV newly diagnosed in the past week<sup>‡</sup> | 84/522 (16.1%) | 25 /183 (13.7%) | 31/181 (17.1%) | 28/158 (17.7%) |
| Median CD4 count (IQR) | 486 (310–658) | 488 (301–668) | 505 (311–670) | 473 (344–640) |
| CD4 count < 350 cells/μL | 181/621 (29.1%) | 68/211 (32.2%) | 61/212 (28.8%) | 52/198 (26.3%) |
| Other Clinical History |  |  |  |  |
| Blood pressure ≥ 140/80 mmHg | | | | |
| No | 2143 (95.4%) | 712 (94.4%) | 709 (96.1%) | 722 (95.6%) |
| Yes | 104 (4.6%) | 42 (5.6%) | 29 (3.9%) | 33 (4.4%) |
| Body Mass Index<sup>**</sup> |  |  |  |  |
| Median (IQR) | 30 (19–41) | 30 (19–41) | 30 (20–40) | 30.1 (20–41) |
| Mid upper arm circumference, cm |  |  |  |  |
| Median (IQR) | 30 (27–34) | 30 (26–34) | 30 (26–33) | 30 (27–34) |

| Hemoglobin Level, g/dl** |  |  |  |  |
| --- | --- | --- | --- | --- |
| Median (IQR) | 12 (11–13) | 12 (11–13) | 12 (11–13) | 12 (11–13) |
| Anemia prevalence by threshold†† |  |  |  |  |
| Haemoglobin < 11 g/dl (WHO definition) | 642/1992 (32.2%) | 206/659 (31.3%) | 210/653 (32.2%) | 226/680 (33.2%) |
| Haemoglobin < 10 g/dl (Other definitions) | 280/1992 (14.1%) | 84/659 (12.8%) | 94/653 (14.4%) | 102/680 (15.0%) |

IQR – Interquartile Range
\*Test-of-Cure visit only performed for One-Time Screening participants who tested positive for an STI at enrollment. Third Trimester visit assessments were routine for all arms, but repeat STI screening with real-time results was provided to Two-Time Screening participants only
†1 ZAR = 0.05 USD
‡ Data collection on smoking and recency of HIV diagnosis (within the prior week) commenced several months after recruitment began; denominators reflect participants who responded.
§Past pregnancy loss history was assessed among participants who had previously been pregnant, and preterm birth history was specific to those with a prior live birth.
¶Standard-of-Care group baseline samples were tested retrospectively for *C. trachomatis*, *N. gonorrhoeae*, and *T. vaginalis* after study completion (April-June 2025); completed for 747 retrievable of 755 stored vaginal swab specimens.
||Participants with a positive rapid syphilis treponemal test result were treated with Benzathine penicillin G.
\*\*Body Mass Index was estimated using weight and height measured at the first antenatal visit (within 27 weeks' gestation).
††The World Health Organization (WHO) defines anemia in pregnancy as a hemoglobin concentration <11 g/dL. We also report on proportion with anemia based on a threshold of 10 g/dL to align with comparator studies, such as the WANTAIM study.

Most characteristics were similar across the study groups (**Table 1**). One exception was a lower proportion reporting prior preterm birth in the Two-Time Screening Group (47/475, 9·9%) compared with the One-Time Screening (59/468, 12·8%) and Standard-of-Care (72/458, 15·9%) groups.

The proportion testing positive for any of the three STIs at baseline was higher in the Standard-of-Care Group (218/747, 29·2%; detected retrospectively with the Gene Xpert in stored vaginal swabs) than in the One-Time (193/754, 25.6%) and Two-Time (186/738, 25·2%) screening groups, in which testing and treatment were conducted in real time. Approximately 86% presenting for antenatal care (**Table 1**) and approximately 80% with infection were asymptomatic (**Figure 2**).

**Figure 2.**
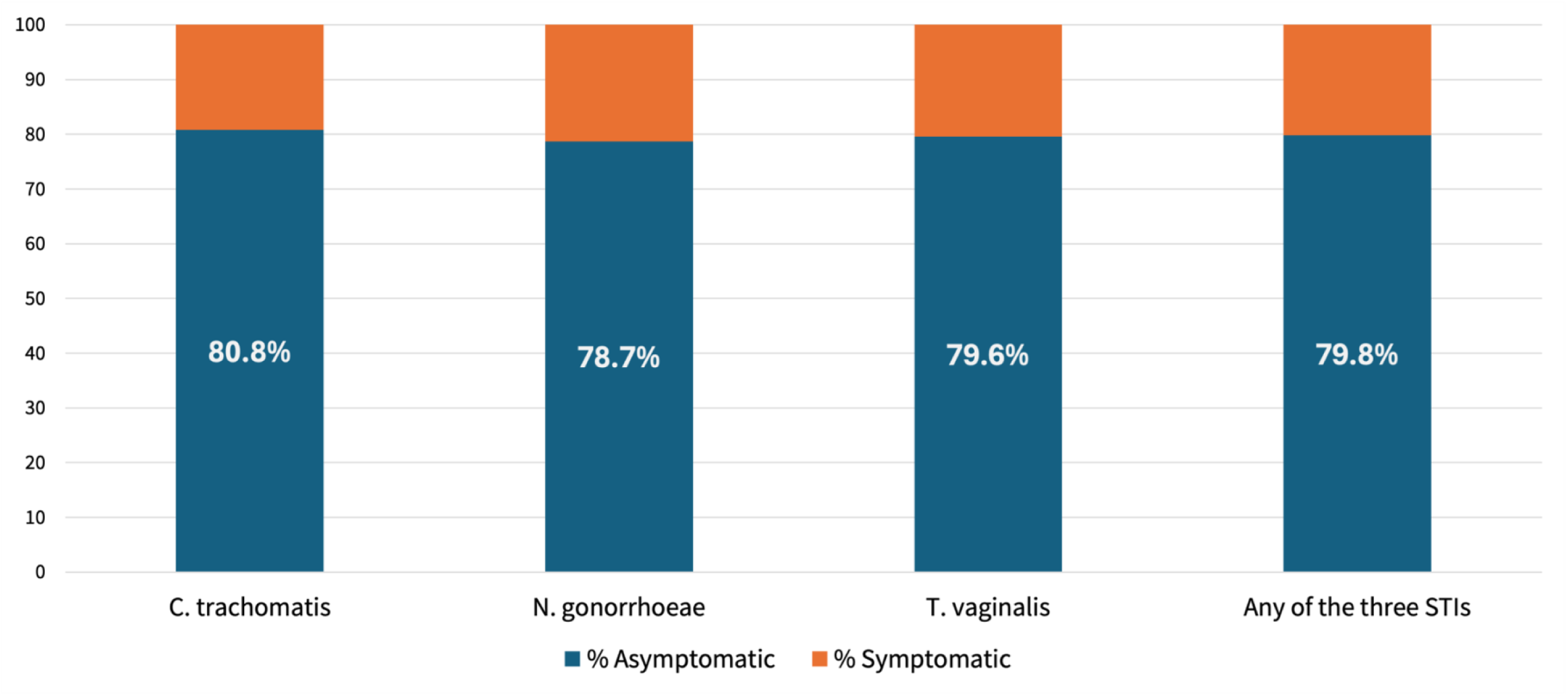
Distribution of asymptomatic and symptomatic laboratory-confirmed STIs at Baseline

### Main Outcomes

Among 1910 participants with live births (**Table 2**), the primary composite outcome of preterm birth or low birthweight occurred in 23·2% of the Standard-of-Care Group, 22·9% of the One-Time Screening Group, and 20·6% of the Two-Time Screening Group. Compared with standard-of-care, the risk ratio (RR) for the composite outcome was 0·99 (95% CI, 0·81–1·21) for One-Time Screening and 0·89 (95% CI, 0.72–1.09) for Two-Time Screening.

**Table 2.**
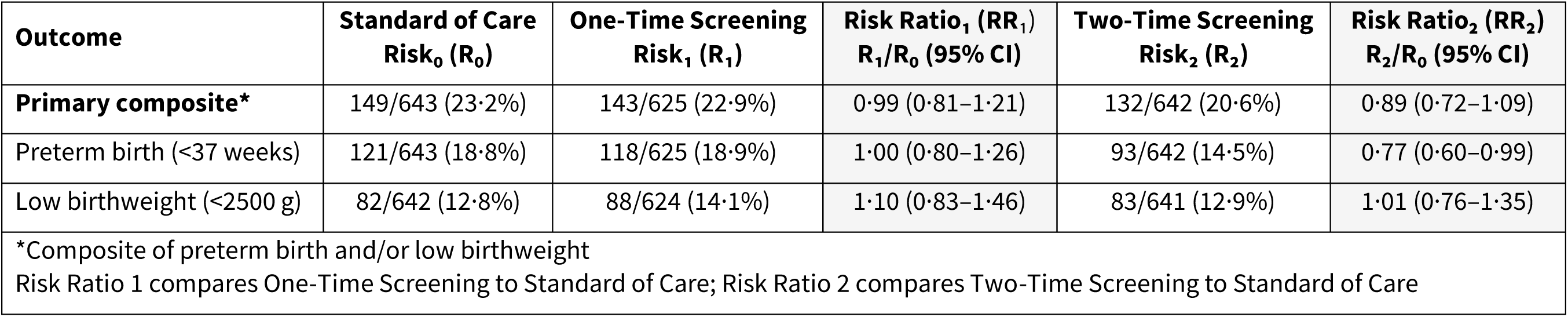
Effect of screening on preterm birth or low birthweight among 1910 participants with live births.

| **Outcome** | **Standard of Care**<br>**Risk<sub>0</sub> (R<sub>0</sub>)** | **One-Time Screening**<br>**Risk<sub>1</sub> (R<sub>1</sub>)** | **Risk Ratio<sub>1</sub> (RR<sub>1</sub>)**<br>**R<sub>1</sub>/R<sub>0</sub> (95% CI)** | **Two-Time Screening**<br>**Risk<sub>2</sub> (R<sub>2</sub>)** | **Risk Ratio<sub>2</sub> (RR<sub>2</sub>)**<br>**R<sub>2</sub>/R<sub>0</sub> (95% CI)** |
| --- | --- | --- | --- | --- | --- |
| **Primary composite\*** | 149/643 (23.2%) | 143/625 (22.9%) | 0.99 (0.81–1.21) | 132/642 (20.6%) | 0.89 (0.72–1.09) |
| Preterm birth (<37 weeks) | 121/643 (18.8%) | 118/625 (18.9%) | 1.00 (0.80–1.26) | 93/642 (14.5%) | 0.77 (0.60–0.99) |
| Low birthweight (<2500 g) | 82/642 (12.8%) | 88/624 (14.1%) | 1.10 (0.83–1.46) | 83/641 (12.9%) | 1.01 (0.76–1.35) |
| *Composite of preterm birth and/or low birthweight<br>Risk Ratio 1 compares One-Time Screening to Standard of Care; Risk Ratio 2 compares Two-Time Screening to Standard of Care |  |  |  |  |  |

Preterm birth occurred in 18·8%, 18·9%, and 14·5% of the Standard-of-Care, One-Time Screening, and Two-Time Screening Groups, respectively (RR 1·00 [95% CI, 0·80–1·26] and 0·77 [95% CI, 0·60–0·99]).

Low birthweight occurred in 12·8%, 14·1%, and 12·9% of the Standard-of-Care, One-Time Screening, and Two-Time Screening Groups, respectively (RR 1·10 [95% CI, 0·83–1·46] and 1·01 [95% CI, 0·76–1·35]).

### Distribution of preterm birth

The distribution of preterm births (**Supplementary Table S1**) showed that the majority were late (34 to <37 weeks), accounting for 84/121 (69·4%) in the Standard-of-Care group, 79/118 (66·9%) in the One-Time Screening group, and 68/93 (73·1%) in the Two-Time Screening group. For early births, extreme preterm births (<28 weeks) accounted for 3/121 (2·5%), 5/118 (4·2%), and 2/93 (2·2%) in the respective study groups, while very preterm births (28 to <32 weeks) occurred in 18/121 (14·9%), 18/118 (15·3%), and 11/93 (11·8%).

### Sensitivity analyses accounting for obstetric history

Although baseline characteristics were generally similar across study groups, the proportion reporting a history of preterm birth among participants who had been pregnant and had a live birth before was lower in the Two-time Screening group (**Table 1**). Consistent with the prespecified analytic plan, we conducted sensitivity analyses to assess the robustness of the secondary observed effect of Two-Time Screening on preterm birth to this imbalance (**Supplementary tables S2A–S2D**). The direction or magnitude of effect were comparable with the main analysis.

### Baseline STI Subgroup

Prespecified exploratory subgroup analyses among participants with laboratory-confirmed STIs at baseline showed generally similar patterns for the primary composite outcome and its components across study group. Subgroup sizes were small and confidence intervals wide (**Supplementary Table S3**).

### Follow up STI testing: Uptake and Positivity

At baseline, treatment uptake among participants diagnosed with an STI was 95% in both intervention groups. Test-of-cure attendance among baseline STI–positive participants in the One-Time Screening Group was 56% (108/193) wherein repeat STI positivity occurred in 18·5% (20/108). At test-of-cure, only 44% (48/108) reported that their partner had sought treatment. In the Two-Time Screening Group, 68% (499/738) attended the third trimester screening (30–34 weeks). Third trimester STI prevalence was 9·2% (46/499); repeat positivity occurring in 20·5% (25/122) of participants who tested positive at baseline, and incident infection in 5·6% (21/377) of those who tested negative at baseline. In all study groups, STI prevalence declined from baseline to the postnatal visit (**Supplementary Table S4**).

### Adverse Events Reporting (Perinatal events)

Among 2,119 participants with pregnancy outcome retrieved, there were 144 (6.8%) miscarriages, 10 (0.5%) ectopic pregnancies, and 8 (0·3%) pregnancy terminations (**Figure 1**). Among all deliveries (N=1957), 47 were stillbirths (2·4%), similarly distributed across study groups. No serious perinatal adverse events, including neonatal hospitalizations, were attributed to the STI screening and treatment intervention (**Table 3**).

**Table 3.**
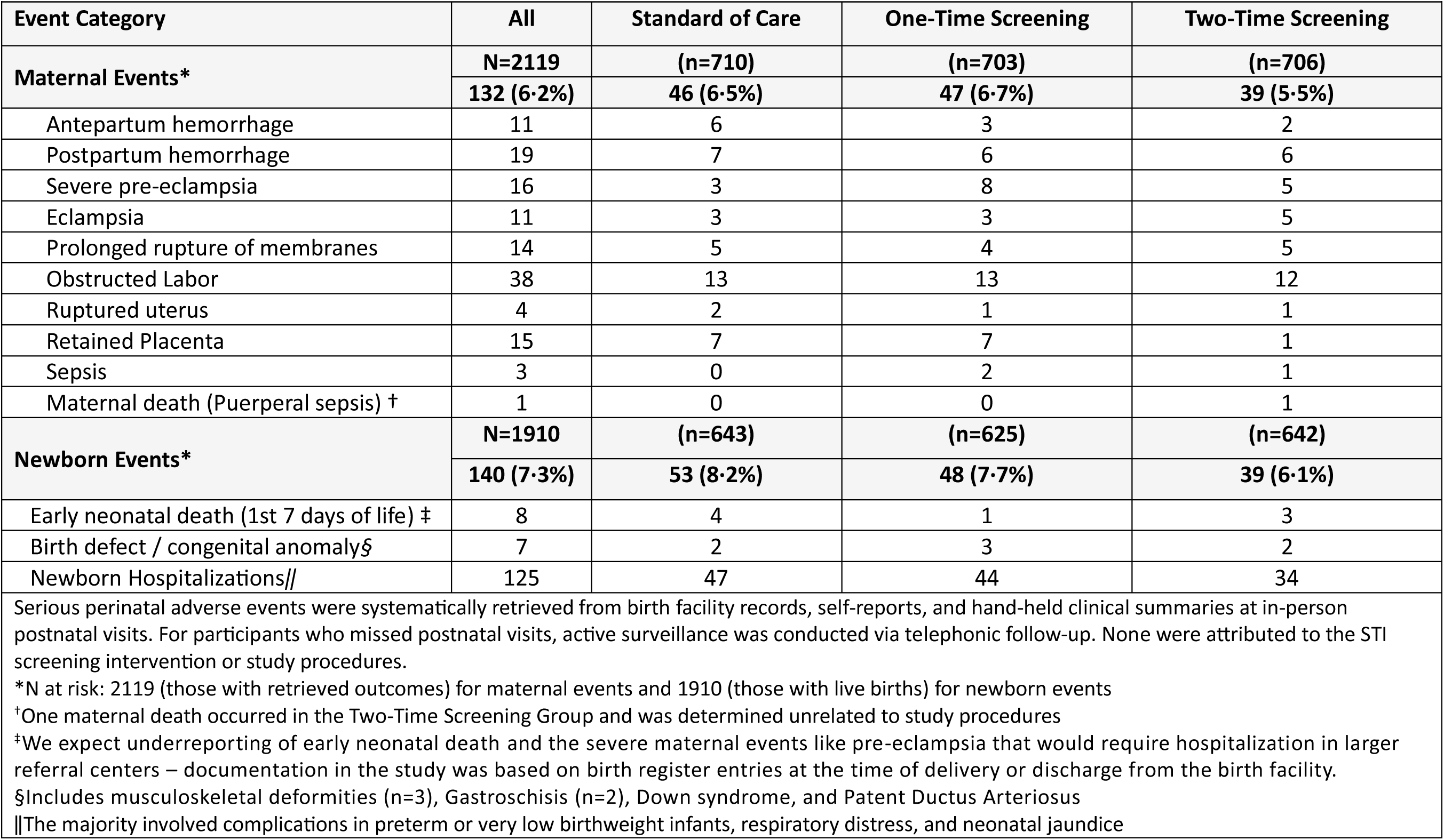
Serious Perinatal Adverse Events.

| **Table 3. Serious Perinatal Adverse Events** |  |  |  |  |
| --- | --- | --- | --- | --- |
| **Event Category** | **All** | **Standard of Care** | **One-Time Screening** | **Two-Time Screening** |
| **Maternal Events\*** | **N=2119** | **(n=710)** | **(n=703)** | **(n=706)** |
|  | **132 (6.2%)** | **46 (6.5%)** | **47 (6.7%)** | **39 (5.5%)** |
| Antepartum hemorrhage | 11 | 6 | 3 | 2 |
| Postpartum hemorrhage | 19 | 7 | 6 | 6 |
| Severe pre-eclampsia | 16 | 3 | 8 | 5 |
| Eclampsia | 11 | 3 | 3 | 5 |
| Prolonged rupture of membranes | 14 | 5 | 4 | 5 |
| Obstructed Labor | 38 | 13 | 13 | 12 |
| Ruptured uterus | 4 | 2 | 1 | 1 |
| Retained Placenta | 15 | 7 | 7 | 1 |
| Sepsis | 3 | 0 | 2 | 1 |
| Maternal death (Puerperal sepsis) † | 1 | 0 | 0 | 1 |
| **Newborn Events\*** | **N=1910** | **(n=643)** | **(n=625)** | **(n=642)** |
|  | **140 (7.3%)** | **53 (8.2%)** | **48 (7.7%)** | **39 (6.1%)** |
| Early neonatal death (1st 7 days of life) ‡ | 8 | 4 | 1 | 3 |
| Birth defect / congenital anomaly§ | 7 | 2 | 3 | 2 |
| Newborn Hospitalizations\|\| | 125 | 47 | 44 | 34 |
| Serious perinatal adverse events were systematically retrieved from birth facility records, self-reports, and hand-held clinical summaries at in-person postnatal visits. For participants who missed postnatal visits, active surveillance was conducted via telephonic follow-up. None were attributed to the STI screening intervention or study procedures.<br>*N at risk: 2119 (those with retrieved outcomes) for maternal events and 1910 (those with live births) for newborn events<br>†One maternal death occurred in the Two-Time Screening Group and was determined unrelated to study procedures<br>‡We expect underreporting of early neonatal death and the severe maternal events like pre-eclampsia that would require hospitalization in larger referral centers – documentation in the study was based on birth register entries at the time of delivery or discharge from the birth facility.<br>§Includes musculoskeletal deformities (n=3), Gastroschisis (n=2), Down syndrome, and Patent Ductus Arteriosus<br>\|\|The majority involved complications in preterm or very low birthweight infants, respiratory distress, and neonatal jaundice |  |  |  |  |

## DISCUSSION

In this three-group randomized trial, antenatal screening and treatment for *C. trachomatis, N. gonorrhoeae, and T. vaginalis* did not significantly reduce the composite primary outcome of preterm birth or low birthweight compared to syndromic management. When the components of the composite outcome were examined separately, neither screening strategy reduced low birthweight. However, two-time screening —once before 27 weeks’ gestation and again between 30- and 34 weeks’ gestation — was associated with a lower risk of preterm birth (23% relative reduction). This pattern persisted in sensitivity analyses accounting for prior preterm birth history. One-time screening with test-of-cure was not associated with a reduction in preterm birth.

The absence of an overall effect on the primary composite outcome of preterm birth or low birthweight is compatible with the WANTAIM trial in Papua New Guinea. WANTAIM evaluated three-time antenatal screening and found no reduction in composite and component outcomes in the full analytic sample.^15^ In Philani, however, two-time screening was associated with a lower risk of the preterm birth component, without corresponding decreased risk of low birthweight. This observed secondary pattern may reflect differences in study populations and epidemiologic context. Preterm birth was more than twice as common in the South African setting than Papua New Guinea, with better nutrition and anemia markers. Substance use patterns may also differentially influence risks related to gestational duration and fetal growth. Philani study participants reported greater alcohol use, while WANTAIM participants used betel nuts, and were more likely to use tobacco.^23^ In the WANTAIM study participants in the control condition received universal malaria prophylaxis with sulfadoxine, an antimicrobial with known anti-chlamydial activity. Malaria is not endemic in the Philani study setting.

The association with lower preterm birth with multiple screenings in the Philani trial is compatible with longstanding observational evidence.^8,9,24^ Ascending infection may trigger inflammatory pathways that promote preterm labor. ^25^ Importantly, the optimal timing of screening remains unclear, though some hypotheses suggest that early screening before 20 weeks’ gestation may be most effective.^26^ In the Philani trial, participants enrolled at a relatively early median of 13 weeks. Repeat testing and treatment may have provided an additional opportunity to address persistent and incident infection in the Two-Time screening group.

Interpretation of early outcomes occurring before the second screening visit (median 30 weeks) requires caution, as these would not have been affected by that later intervention. Notably, the distribution of preterm births showed a higher proportion of late preterm births (34 to <37 weeks) in the Two-Time Screening group (73·1%) relative to the One-Time (66·9%) and Standard-of-Care (69·4%) groups. This exploratory pattern suggests the second intervention may have delayed delivery in some participants, shifting outcomes toward less severe prematurity—a hypothesis requiring confirmation in future trials.

The risk of low birthweight was similar in the intervention and control groups. Low birthweight reflects both gestational duration and fetal growth and may be less sensitive to these interventions. Mechanisms linking genital infections to fetal growth restriction are less clearly defined. Inflammation may impair deep placentation or signaling for insulin-like growth factor 1 and growth hormone,^27,28^ and antimicrobial treatment alone may be insufficient to reverse effects once underway. A more detailed understanding of how STIs affect fetal growth, or how antibiotics mitigate this risk is needed. Integrated approaches to reduce adverse birth outcomes that address overlapping infectious, inflammatory, and nutritional contributors may be needed to optimize the conditions for a healthy pregnancy. ^29^.

WANTAIM observed a significant reduction in the primary composite outcome of preterm birth or low birthweight among women with *N. gonorrhoeae* at baseline. In Philani, analogous subgroup analyses among participants with baseline STIs showed directionally similar patterns for two-time screening but were underpowered for statistical inference.

Our trial had limitations. The composite outcome, selected a priori to align with similar trials, may have limited sensitivity. Future trials should consider preterm birth and growth-restricted outcomes independently. Subgroup analyses in participants with STI were exploratory underpowered for definitive inference, and antenatal infection patterns—such as persistence or reinfection—could not be compared between study groups. We did not test and treat other genital tract conditions, such as bacterial vaginosis or *Mycoplasma genitalium* infection though randomization likely distributed these evenly. As a single-site trial, generalizability may be limited where other drivers of preterm birth, such as malaria, malnutrition, or non-infectious etiologies, predominate. Baseline STI prevalence was higher in the Standard-of-Care Group. In this group, specimens were stored in GeneXpert transport media for a variable duration of 6 months to 4 years depending on time of collection and tested retrospectively after trial completion, whereas screening groups were tested in real time. The extent to which storage conditions may have contributed to this difference is unclear. We did not adjust for baseline STI status in the primary models; however, prespecified STI subgroup analyses are presented for descriptive and exploratory comparison. The observed decline in postnatal STI prevalence in the Standard-of-Care group may reflect a combination of syndromic treatment of symptomatic infections throughout the antenatal period, interim care received outside the study, and spontaneous clearance of some infections. Strengths of the study include individual randomization, a rigorous yet pragmatic design that increases the potential for real-world implementation, and the use of gold standard point-of-care diagnostics. Maternal HIV-related health was also comparable across study groups at baseline and aligned with regional trends, suggesting generalizability to women living with HIV in similar high-prevalence settings. ^30,31^. As a pragmatic hybrid effectiveness trial, only a little over half attended the test of cure, which was a non-routine antenatal visit. Attendance at the routine third-trimester screen (68%), on the other hand, was limited by participants with early pregnancy loss or delivery before the scheduled 30-week visit. All participants, regardless of follow-up were included in the modified intention-to-treat analysis to preserve randomization.

As of 2017, only 14 countries globally had adopted national antenatal screening policies for *C. trachomatis or N. gonorrhoeae*.^32^ In light of the observed findings, including the absence of an effect on the primary composite outcome and a secondary association with preterm birth, further evaluation of antenatal STI screening strategies with respect to timing and frequency remains necessary. The findings may also be informative for higher-income settings, such as the United States, where preterm birth rates are still one of the highest in the world, and where these curable STIs often go undiagnosed near delivery.^33^ Together, WANTAIM, Philani, and forthcoming trials^34,35^ are building the evidence to determine whether, how, and for what populations antenatal STI screening can effectively reduce adverse birth outcomes.

## CONTRIBUTORS

JDK and AMM conceived and designed the study (Conceptualization, Methodology). RPH was site investigator and oversaw implementation (Investigation, Supervision), with MMM as research manager responsible for daily site operations and project administration. RMG supported site efforts. CMB coordinated study fidelity and troubleshooting, monitoring of data quality, and facilitated independent trial oversight by the Data and Safety Monitoring Board (Project administration, Supervision, Validation). FM managed the study database and supported data cleaning (Data curation, Software), under review by CMB and MW. MW served as lead statistician, performed the primary analyses independently of CMB (Formal analysis, Validation), and guided reporting. HJ provided laboratory and data support, including microbiological specimen banking and indexing (Data curation, Resources, Validation). JDK, AMM, and CMB drafted the initial manuscript (Writing – original draft). CAM, CMT, and NL contributed to study oversight, supported development of secondary study objectives (Resources, Supervision), and reviewed and revised the manuscript (Writing – review & editing). All authors reviewed and approved the final manuscript. FM, MM, CMB, and MW, had full access to the study data, verified the underlying data, and take responsibility for the decision to submit for publication.

## Data Availability

All data produced in the present study are available upon reasonable request to the authors

## ACKNOWLEDGMENTS

We thank the individuals who participated in the Philani Ndiphile Trial, and communities who supported them; the local trial staff; and the Data and Safety Monitoring Board (Joseph D. Tucker, Chelsea Morroni, Mary-Ann Davis, Trevor Pickering, and Katherine Gill). Cepheid (Sunnyvale, CA, USA) loaned the Gene Xpert diagnostic equipment for each clinic sites and supported consumables at subsidized cost.

---

**Supplementary Table S1.** Preterm Births by Gestational Age at Delivery.

| Preterm Birth Category* | All Preterm | Standard of Care | One-Time Screening | Two-Time Screening |
| --- | --- | --- | --- | --- |
|  | N = 332 | n = 121 | n = 118 | n = 93 |
| Extremely Preterm (< 28 weeks) | 10 (3·0%) | 3 (2·5%) | 5 (4·2%) | 2 (2·2%) |
| Very Preterm (28 to <32 weeks) | 48 (14·4%) | 18 (14·9%) | 18 (15·3%) | 11 (11·8%) |
| Moderately Preterm (32 to < 34weeks) | 44 (13·3%) | 16 (13·2%) | 16 (13·6%) | 12 (12·9%) |
| Late Preterm (34 to <37 weeks) | 230 (69·3%) | 84 (69·4%) | 79 (66·9) | 68 (73·1%) |
| *World Health Organization Classification |  |  |  |  |

**Supplementary Table S2.**
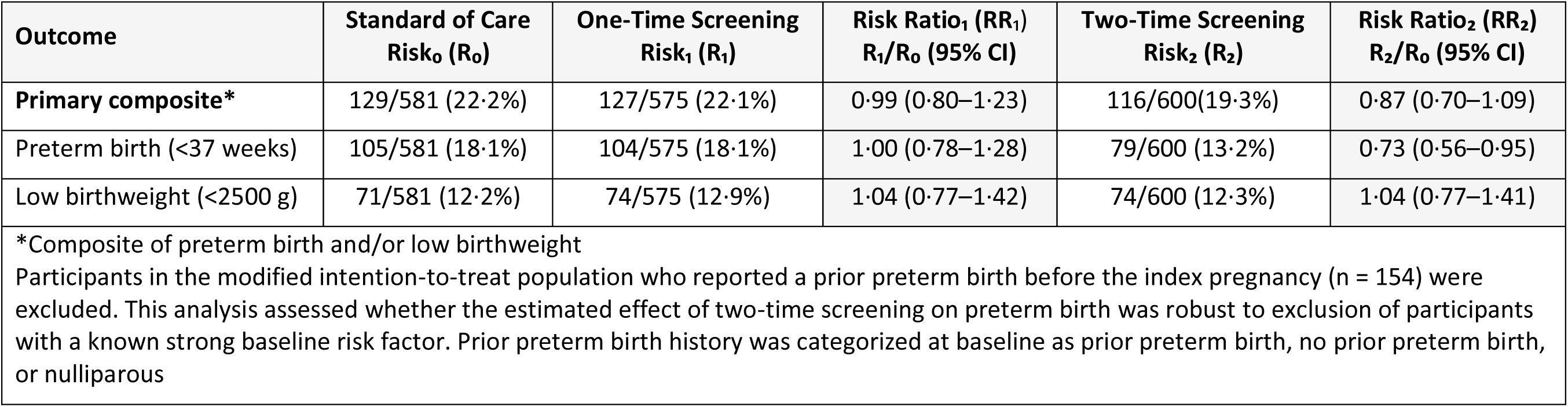
A. Sensitivity analysis excluding participants with a history of preterm birth at baseline (N=1756)

**Supplementary Table S2.**
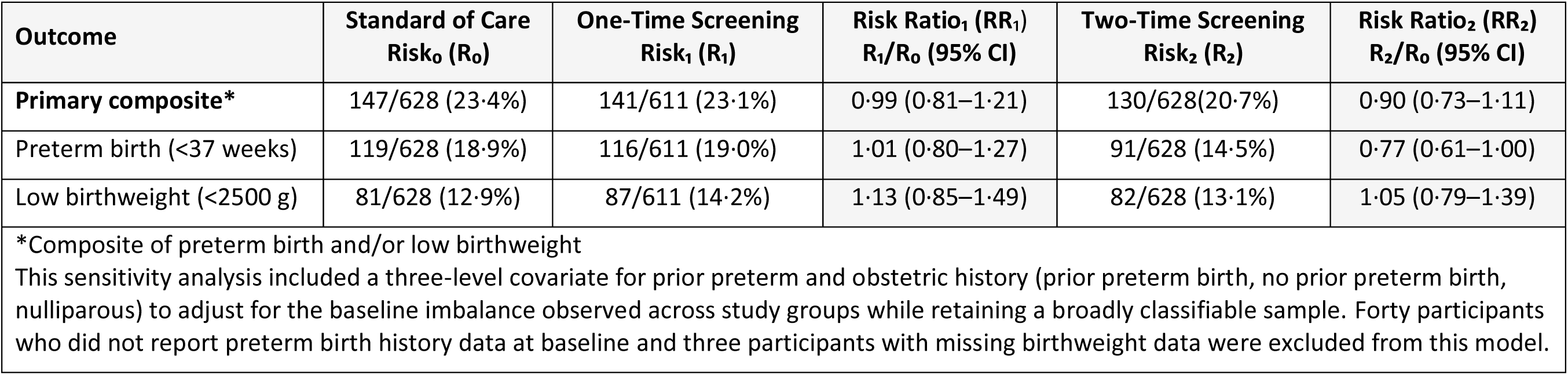
B. Sensitivity analysis adjusting for prior obstetric history as a covariate in the model (N=1867)

**Supplementary Table S2.**
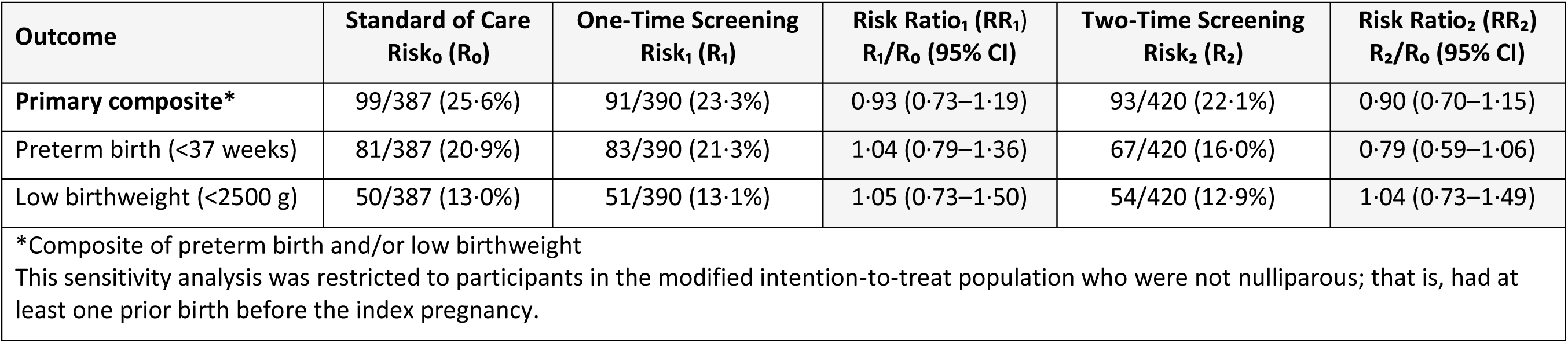
C. Sensitivity analysis excluding nulliparous participants at baseline (N = 1197)

**Supplementary Table S2.**
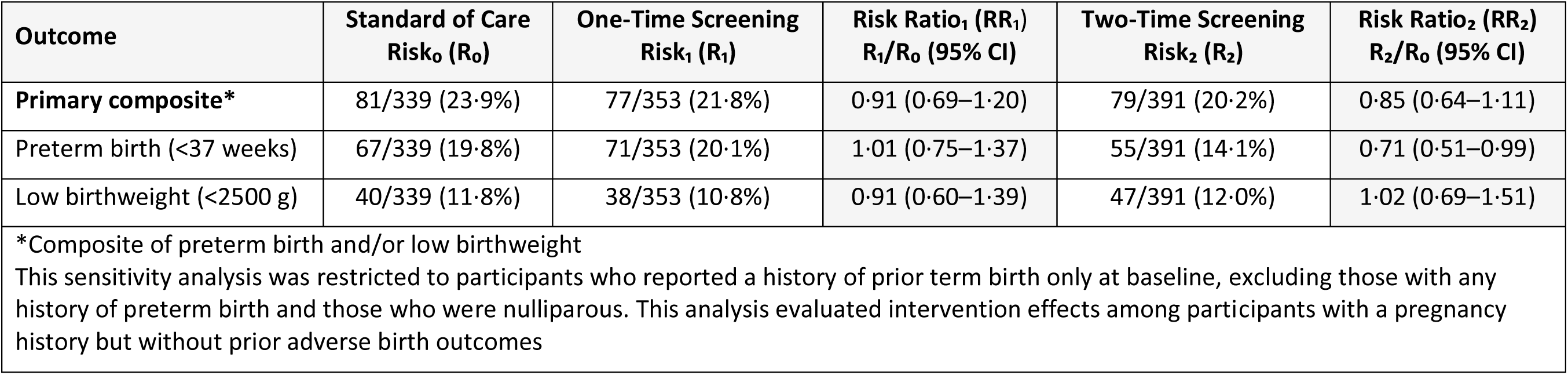
D. Sensitivity analysis restricting to participants with prior term births (N = 1083)

**Table S3.** Subgroup analyses of intervention effect in participants with *C. trachomatis*, *N. gonorrhoeae*, and *T. vaginalis* at baseline.

|  | Standard<br>of Care | One-Time Screening | Risk Ratio 1<br>(95% CI) | Two-Time Screening | Risk Ratio 2<br>(95% CI) |
| --- | --- | --- | --- | --- | --- |
|  | (R<sub>0</sub>) | (R<sub>1</sub>) | (R<sub>1</sub>) / (R<sub>0</sub>) | (R<sub>2</sub>) | (R<sub>2</sub>) / (R<sub>0</sub>) |
| ***C. trachomatis* (N = 297)** | **n = 113** | **n = 90** |  | **n = 94** |  |
| Primary composite* | 27 (23.9%) | 20 (22.2%) | 0.93 (0.56–1.55) | 21 (22.3%) | 0.93 (0.57–1.54) |
| Preterm birth | 21 (18.6%) | 17 (18.9%) | 1.02 (0.57–1.81) | 14 (14.9%) | 0.80 (0.43–1.49) |
| Low birth weight | 16 (14.2%) | 14 (15.6%) | 1.10 (0.57–2.13) | 12 (12.8%) | 0.90 (0.45–1.81) |
| ***N. gonorrhoeae* (N = 110)** | **n = 44** | **n = 32** |  | **n = 34** |  |
| Primary composite* | 7 (19.4%) | 9 (31.0%) | 1.77 (0.74–4.25) | 4 (15.4%) | 0.74 (0.24–2.32) |
| Preterm birth | 7 (15.9%) | 6 (18.8%) | 1.18 (0.44–3.17) | 3 (8.8%) | 0.55 (0.15–1.99) |
| Low birth weight | 2 (4.5%) | 7 (21.9%) | 4.81 (1.07–21.65) | 2 (5.9%) | 1.29 (0.19–8.72) |
| ***T. vaginalis* (N = 201)** | **n = 67** | **n = 70** |  | **n = 64** |  |
| Primary composite* | 19 (28.4%) | 20 (28.6%) | 1.01 (0.59–1.71) | 11 (17.2%) | 0.61 (0.31–1.17) |
| Preterm birth | 14 (20.9%) | 18 (25.7%) | 1.23 (0.67–2.27) | 9 (14.1%) | 0.67 (0.31–1.44) |
| Low birth weight | 12 (17.9%) | 12 (17.1%) | 0.96 (0.46–1.98) | 7 (10.9%) | 0.61 (0.26–1.45) |
| **Any of the three STIs (N = 507)** | **n = 180** | **n = 162** |  | **n = 165** |  |
| Primary composite* | 46 (25.6%) | 41 (25.3%) | 0.99 (0.69–1.42) | 31 (18.8%) | 0.74 (0.49–1.10) |
| Preterm birth | 36 (20.0%) | 34 (21.0%) | 1.05 (0.69–1.59) | 21 (12.7%) | 0.64 (0.39–1.04) |
| Low birth weight | 26 (14.4%) | 28 (17.3%) | 1.20 (0.73–1.95) | 17 (10.3%) | 0.71 (0.40–1.27) |

\*Composite of preterm birth and/or low birthweight

**Table S4.** STI Positivity at Baseline, Third Trimester, and Postnatal Visits, by Study Group.

|  |  |  |  |
| --- | --- | --- | --- |
|  | Baseline (<27 weeks) | Third Trimester (30-34 weeks) | Postnatal (< 2 weeks after delivery) |
| One Time Screening | n = 754 |  | n = 296 |
| C. trachomatis | 112 (14.9%) | Not tested | 14 (4.7%) |
| N. gonorrhoeae | 37 (4.9%) |  | 8 (2.7%) |
| T. vaginalis | 81 (10.7%) |  | 8 (2.7%) |
| Any of the three STIs | 193 (25.6%) |  | 25 (8.4%) |
| Two Time Screening | n = 738 | n=499 | n = 328 |
| C. trachomatis | 107 (14.5%) | 24 (4.8%) | 8 (2.4%) |
| N. gonorrhoeae | 36 (4.9%) | 13 (2.6%) | 4 (1.2%) |
| T. vaginalis | 71 (9.6%) | 14 (2.8%) | 9 (2.7%) |
| Any of the three STIs | 186(25.2%) | 46 (9.2%) | 21 (6.4%) |
| Standard of Care | n = 747 |  | n = 313 |
| C. trachomatis | 134 (17.9%) | Not tested | 20 (6.4%) |
| N. gonorrhoeae | 50 (6.7%) |  | 10 (3.2%) |
| T. vaginalis | 83 (11.1%) |  | 19 (6.1%) |
| Any of the three STIs | 218 (29.2%) |  | 41 (13.1%) |
| *Baseline: enrollment at the first antenatal visit (before 27 completed weeks of gestation). Samples from participants in the Standard-of-Care Group were collected in GeneXpert transport media and tested after study completion. |  |  |  |
| †Third Trimester: Visit occurred between 30 and 34 weeks of gestation. Only participants in the Two-Time Screening Group were tested in real time; samples from the One-Time Screening and Standard-of-Care were stored and are pending testing. |  |  |  |
| ‡Postnatal: Participants who attended a visit within 2 weeks of delivery were tested in all study groups |  |  |  |

